# CovTransformer: A transformer model for SARS-CoV-2 lineage frequency forecasting

**DOI:** 10.1101/2024.04.01.24305089

**Authors:** Yinan Feng, Emma E. Goldberg, Michael Kupperman, Xitong Zhang, Youzuo Lin, Ruian Ke

## Abstract

With hundreds of SARS-CoV-2 lineages circulating in the global population, there is an urgent need for forecasting lineage frequencies and thus identifying rapidly expanding lineages. To address this need, we constructed a framework for SARS-CoV-2 lineage frequency forecasting (CovTransformer), based on the transformer architecture. We designed our framework to navigate challenges such as a limited amount of data with high levels of noise and bias. We first trained and tested the model using data from the UK and the US, and then tested the generalization ability of the model on data collected across the globe. Remarkably, the model makes predictions two months into the future with high levels of accuracy in 31 countries. Finally, we show that our model performed substantially better than the current gold-standard, i.e. a regression-based model implemented in Nextstrain. Overall, our work demonstrates transformer models represent a promising approach for lineage forecasting and pandemic monitoring.

## Introduction

SARS-CoV-2 is continuously evolving new variants that increase transmission fitness and/or evade population immunity (Carabelli et al. 2023; Markov et al. 2023; Meijers et al. 2023; Volz 2023). As a result, many waves of infection around the world were caused by variants of concern (such as subvariants of the Delta and the Omicron lineages), leading to large numbers of infections and a high death toll (Dong et al. 2020; Ritchie et al. 2020). Vaccination is an important and effective tool to reduce the level of transmission along with morbidity and mortality (Polack et al. 2020; Baden et al. 2021). However, with the frequent origination of immune escape variants (Harvey et al. 2021; Rössler et al. 2023; Wilks et al. 2023), there is a continued need for predicting the rate and magnitude of spread for new variants that emerge in a population. This has become increasingly important especially with dozens or hundreds of minor variants circulating and competing in the global population (Beesley et al. 2023). Accurate prediction would allow for timely formulation of new vaccines that target the future dominant variants, and for more focused experimental efforts to understand the pathogenicity and the molecular characterization of the extent of immune escape of variants that have high potential to spread (Stockdale et al. 2022).

Existing forecasting tools have focused on predicting future trajectories of the numbers of COVID-19 cases and deaths (Du et al. 2023; Grubaugh et al. 2019; Hub 2024). However, a predictive tool focusing on forecasting dynamics of variant frequencies is still lacking. The potential for spread of a variant when it first emerges has been predicted based on the extent of structural change on the spike protein leading to immune escape (Harvey et al. 2021; Chen et al. 2022), but the potential for an immune escape mutant to efficiently transmit in the population is unknown. Regression or mechanistic models predict the fitness of a lineage and thus the potential for spread from time series of variant frequency data (Volz 2023; Beesley et al. 2023; Nextstrain 2023; Abousamra et al. 2023). However, due to the noise arising from data collection and deposition (as we show below), this general approach suffers from large uncertainty in prediction. We previously found that robust and accurate predictions require several weeks of data from multiple countries (van Dorp et al. 2021).

In this work, we constructed a transformer model (Vaswani et al. 2017; Dosovitskiy et al. 2021), called Cov-Transformer, to forecast the future frequency of existing SARS-CoV-2 lineages from noisy lineage frequency time series data. Machine learning approaches have been successfully applied to time series analysis and various problems in COVID-19 pandemic response (Che et al. 2018; Song et al. 2018; Syrowatka et al. 2021). In particular, state-of-the-art transformers (Vaswani et al. 2017), distinct for their self-attention mechanism, are renowned for their exceptional ability to capture complex sequential patterns and long-term relationships. This enables the nuanced detection of intricate patterns and long-term dependencies within data sequences, achieving success in many problems such as natural language processing (Vaswani et al. 2017; Brown et al. 2020; Devlin et al. 2018), computer vision (Dosovitskiy et al. 2021; Khan et al. 2022; Kirillov et al. 2023; Girdhar et al. 2019), multimodal learning (Xu et al. 2023; Radford et al. 2021), and time series analysis (Wen et al. 2023; Zhang & Yan 2022; Gao et al. 2022; Song et al. 2018). Our approach integrates transformers within a broader strategy to address the complex task of forecasting pandemic lineage frequencies. This methodology is not limited to the application of advanced machine learning techniques; it systematically tackles inherent challenges such as large noises, reporting delays, input biases, variable label quality, and the scarcity of comprehensive data across a wide array of lineages. We set a precedent for utilizing machine learning tools in lineage-level frequency forecasting, demonstrating a tailored solution to the unique obstacles presented by pandemic data analysis. Here, we first show that simple forecasting of emerging variant frequencies based on regression leads to erroneous predictions due to the inherently noisy nature of the data. Then we demonstrate that our machine learning transformer model accurately forecasts lineage frequencies and, importantly, identifies lineages that eventually reach high frequencies from a collection of newly emerged lineages.

## Results

### Data collection and a regression approach using recent variant frequency time series

To predict variant frequency trajectories, we used the metadata for SARS-CoV-2 sequences from GISAID (Elbe & Buckland-Merrett 2017, downloaded on Feb. 26, 2024, see Methods for details). For each viral sequence, we noted both the collection date when a viral sample was collected from an infected individual, and the reporting date when the sequence became available in the database. In general, there is a wide distribution of the delay in reporting, defined as the difference in days between the collection date and the reporting date (Fig. S1). The mean and median delays in reporting are 30.1 and 17 days, respectively, whereas the standard deviation is 51.6 days. As we show below, the long reporting delays leads to large uncertainties and biases in lineage frequency time series especially for days immediately before the date of access to the database (see Fig. S2 for example).

To form a dataset for training our models below, we first constructed an input dataset where we assumed the database was accessed on each day between Jan. 1, 2020 and Feb. 22, 2024. For each day of access, we calculated the frequency time series of each Pango lineage for the past two months by only considering the viral genetic sequences available in the database before or on the access date. Overall, there are a total of 145,419 time series for two countries’ data (US and UK), which serve as an input dataset for our models.

To construct a target dataset for model evaluation and testing, we calculated the frequency of each Pango lineage (Rambaut et al. 2020) on each day, using all the sequences available when we downloaded the data, i.e. an access date of Feb. 22, 2024. We termed these the ‘final’ frequencies. For the ground truth frequencies used as targets to train and test the model, we fit a smoothing spline to the final frequencies to remove the inherent noise from the data due to day-to-day variations in sampling.

One simple forecasting approach is to estimate the growth rate of a lineage using regression on recent lineage frequencies, and then project future lineage frequency based on the estimated growth rate. We found that because of the reporting delay, the Pango frequencies within two weeks’ time of the access date were often substantially different than the final frequencies, and typically there was no viral genome reported within 1–3 days prior to the access date (Fig. S2). See Fig. 1a for an example, where the frequency time series of XBB.1.9.1 in the US collected on the access date *A* (shown as red dots) are used as model inputs. The frequencies on the access date are very different from the final frequencies, especially at time points close to the access date. This high level of uncertainty in the lineage frequency time series due to reporting delay makes the simple regression approach highly unreliable (Fig. S2). Nonetheless, we used the prediction from this approach as a baseline model below for our machine learning model to compete against.

**Figure 1.**
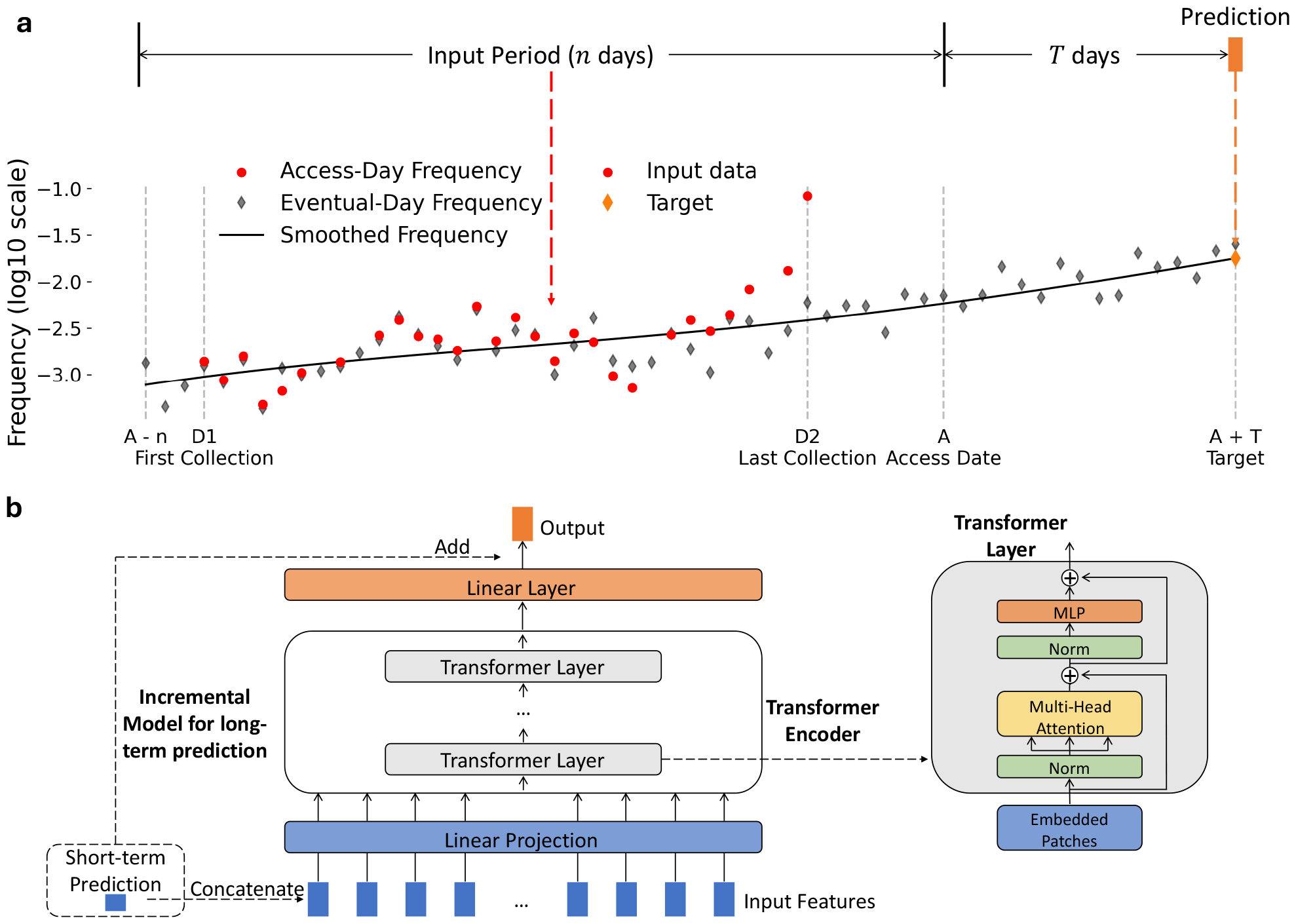
Schematics of model input, output and architecture. **(a)** An example of input and target data for the transformer model. Shown is the time series data for the lineage XBB.1.9.1 in the US (accessed on Feb. 23, 2023). The Input Period is the period (*n*=42 days before the access date *A*) when lineage frequency time series were collected. Red dots show Access-Day Frequency, representing the time series based on data accessed on date *A*, serving as input for our models. The model makes predictions of the lineage frequency *T* days after the access date. The error of the model is calculated as the difference (in log10) between model-predicted frequency and the target frequency, which is calculated from a smoothing spline (black line) through the frequencies eventually measured (gray dots, based on all data collected by Feb. 26, 2024). **(b)** Model architecture. We employed a transformer with a linear input layer preceding it and a linear output layer following it. The transformer layer contains a multi-head self-attention layer, followed by an MLP. We used an incremental method to enhance long-term predictions by including short-term (i.e. 14-day) predictions, as shown in the dotted line part.

### Development of a transformer model

We developed a transformer model to forecast future frequencies of each Pango lineage (see Methods for details). The transformer at its core, is characterized by its distinctive attention mechanism, known as multi-head self-attention. This mechanism enables the model to weigh the significance of each element in a time series against all others, enabling it to capture intricate relationships and long-term dependencies within data sequences, whether in text, time series, or other temporal data (Vaswani et al. 2017; Xu et al. 2023; Wen et al. 2023).

In brief, our model employs a shallow transformer network with a linear input layer preceding it for patch embedding and a linear output layer. The model architecture is illustrated in Fig. 1b. It uses the frequency time series of a Pango lineage in a country as inputs to predict the frequencies of the lineage at a future date (14, 21, 28, 35, 42 and 60 days into the future). We employed an incremental approach to enhance long-term model prediction. More specifically, we developed 6 model variants to make predictions on the 6 future dates. The first model variant is trained to forecast on day 14, and then the 14-day predictions were then concatenated with the input feature as inputs for other 5 model variants. This approach combines short-term insights with long-term forecasting, resulting in improved accuracy. Note that the model at this stage concerns predictions for a single lineage in one country only. It does not consider predictions from other extant lineages at the same time period, although we include this consideration later on.

The error of the model is calculated as the mean absolute error (MAE) between the log10 of the model predicted frequency and the log10 of the target frequency. We calculate the error on the log10 transform of the frequencies because in general, the size of each of the lineages increases or decreases exponentially. One characteristic of the dataset is that there are only a few lineages that rose to a high frequency to become the dominant lineage in the population, while most lineages stayed at relatively low frequencies (Fig. 2b). To address this issue of data imbalance, we applied an exponential function to weight the loss function during model training, such that the errors between target and model prediction from lineages that rose to a high frequency were weighted heavily in the training. In this way, our model is trained to better identify lineages that will become dominant when their frequencies are still low (Fig. 2b).

**Figure 2.**
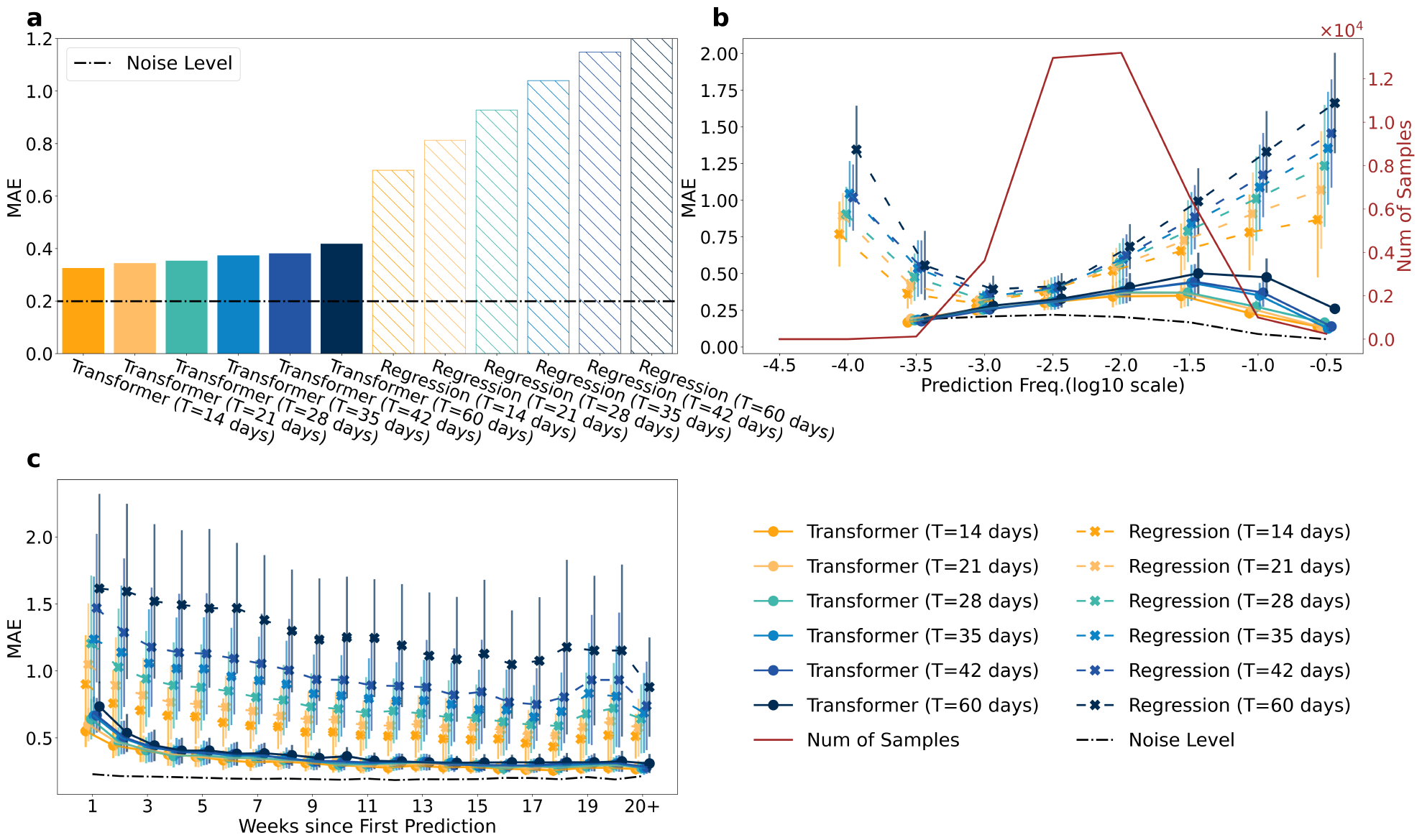
The transformer model outperforms linear regression models on data from UK and US across evaluation categories. In all panels, the noise level (dashed line) is calculated as the MAE between the smoothed final frequency and the final frequency (on a log10 scale). **(a)** Overall model performance of the transformer model versus the regression model for six different future days. **(b)** MAE categorized by the model-predicted frequencies. Results were binned by each 0.5 interval on the log10 of the frequencies. The transformer model performed substantially better than the regression model especially for high frequency predictions: it identifies lineages that would rise to a high frequency much better than the regression model. The number of samples (red line) indicates the number of total valid input time series in each bin. **(c)** MAE categorized by the week since the first prediction day for each lineage (the first day when there are 14 non-zero frequencies in the model input). The transformer has superior performance compared to the regression model, especially in the initial weeks following the emergence of a new variant, when data is scarce.

### The transformer-based model performs well in various evaluation categories

We first trained and tested our model using data derived from the UK and the US, where most SARS-CoV-2 genomic sequences have been collected. We split the dataset containing all lineages such that the data before Jan. 1, 2023 were used as the training and validation dataset and the data between Jan. 1, 2023 and Jan. 1, 2024 were used as the testing dataset. This leads to 107,712 training data and 37,707 testing data.

We tested all the model variants and compared their performance with model predictions from the simple regression approach mentioned above. As shown in Fig. 2a, our model outperforms the regression model across all prediction time points substantially. This performance advantage is especially pronounced in long-term forecasts (e.g. 60 days into the future) where the regression models exhibit escalating error rates. In contrast, the transformer models maintain a lower and more stable loss growth, indicating robustness in handling extended prediction horizons. Remarkably, the MAE for the transformer model predictions ranges between 0.32 and 0.42 for the 6 future dates. These MAEs are only slightly above the mean noise level of the data, i.e. 0.2, calculated as the mean difference between the actual final frequencies derived from the sequence data and the smoothed target frequencies. This means we expect the average difference between our predictions and the ground truth frequencies to be approximately 0.42 on a log10 scale (i.e. 2.6 fold difference on a linear scale) even for predictions 2 months into the future.

We next examined the MAEs using two different categorizations. First, we derived statistics of MAEs according to the predicted frequency of the lineage (Fig. 2b). Our models show remarkable consistency across various prediction frequencies, even though the number of training datasets is very low when the predicted frequency is high. This is critical for lineage surveillance because of the importance of accurate and early identification of lineages that eventually rise to a high frequency in the population. The regression models, in contrast, exhibit pronounced inaccuracies and false positive rates, particularly in these categories. Second, we derived statistics of MAEs according to the time since our model was able to make the first prediction, i.e. defined as the day when a lineage has at least 14 days of non-zero data points in the past 42 days. Fig. 2c shows that even when the number of input data is low, i.e. a few weeks after the lineage appeared in the database, our model makes accurate predictions, whereas the regression model performed poorly.

### Implementing the models for real-time forecasting shows they accurately predict lineage trends in the US and the UK

The results above showed that the transformer models perform well in making predictions for individual lineages. However, in practice, dozens of lineages may coexist in a population at the same time. Therefore, when implementing our models to deal with real-time data, we normalized the predictions of individual lineages such that the sum of all extant lineage frequencies is 1. Fig. 3 shows comparisons between the raw data and our model predictions for the US and the UK during the period between Nov. 30, 2022 and Feb. 18, 2024. Model predictions shown on each day were made using lineage frequency time series collected for an access date 28 or 60 days prior to the day of prediction. The model predictions agree well with the raw data. In particular, our models correctly predicted the rise of XBB lineages in the first half of 2023 and the JN lineages in late 2023, emphasizing their utility in making accurate real-time forecasts and identifying highly transmissible SARS-CoV-2 lineages.

**Figure 3.**
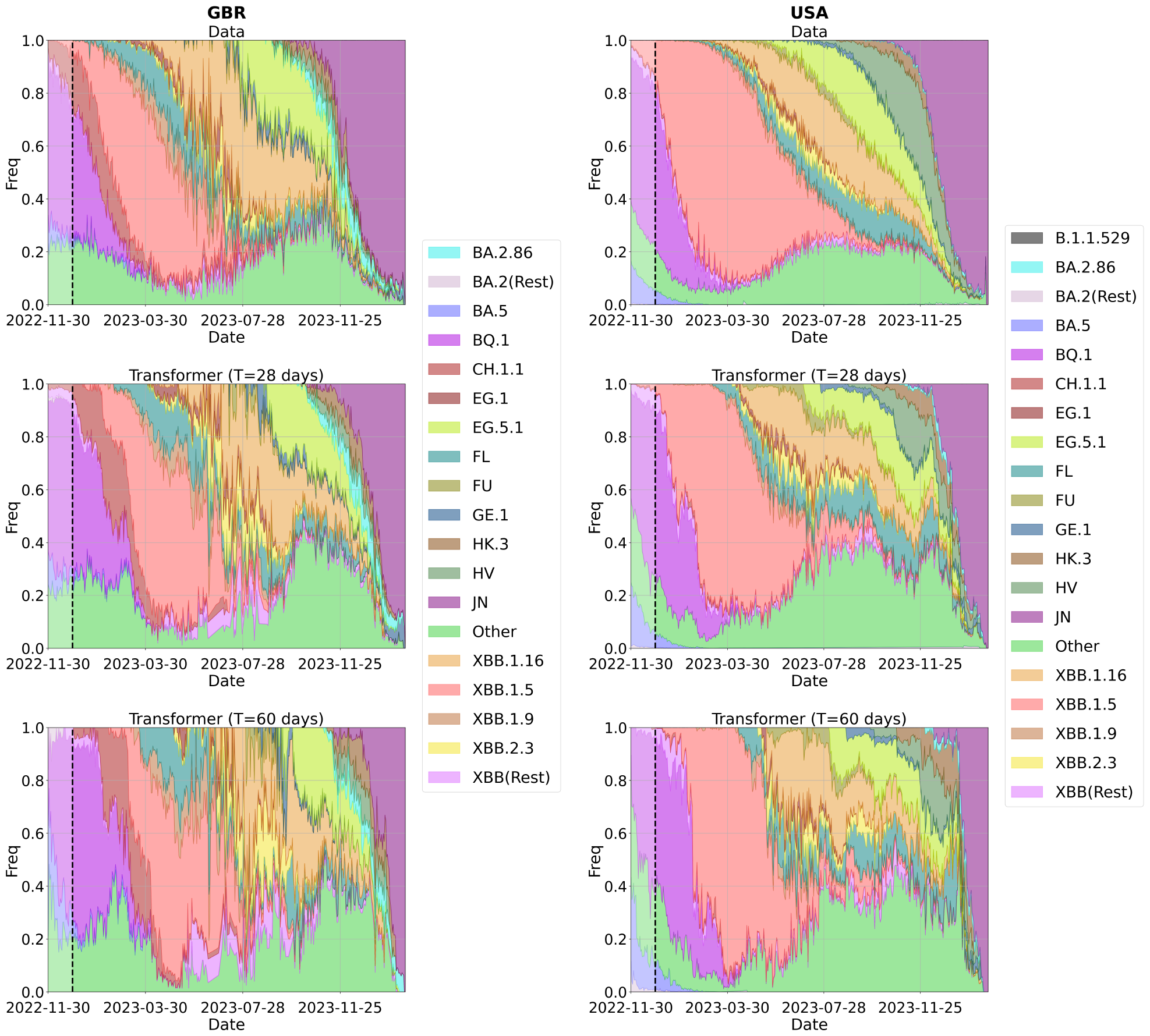
Retrospective predictions from the transformer model agree well with raw data from the UK and the US. Upper panels show the lineage frequencies over time derived from the raw data. Middle and lower panels show the frequencies from the 28 and 60 day predictions from the transformer model. The model-predicted frequencies on a day were calculated by first applying the transformer model to time series of each extant lineage assuming an access date of 28 or 60 days prior to the prediction date, and then normalizing the predictions for all lineages such that the sum of frequencies is 1 on each day. The lineages were clustered and color coded based on Nextstrain clades only for the purpose of visualization. The dashed line denote the date (Dec. 31, 2022) after which the raw data was not used for model training.

### Model generalization and ablation test

Given the highly accurate performance of our models on the data sets from the US and the UK, we further applied these trained models to data from over 100 different countries not used in training, to assess the generalization ability of our models to frequency data from countries with lower genomic surveillance intensity. Our model performance is surprisingly robust for countries with relatively high numbers of available genomic sequences (Fig. 4). For example, for 31 countries (each with more than 20,000 sequences reported), the MAEs of our model predictions are below 0.5. Moreover, 24 of 27 countries with more than 70,000 sequences reported have MAEs below 0.5, and the MAEs of the other three countries are close to 0.5. In addition, we found a clear correlation between the intensity of a country’s sequencing efforts and the performance of our model, with a Pearson correlation coefficient of -0.82 (Fig. 4). This correlation underscores the importance of comprehensive sequencing in enhancing the precision of predictive models in viral genomics.

**Figure 4.**
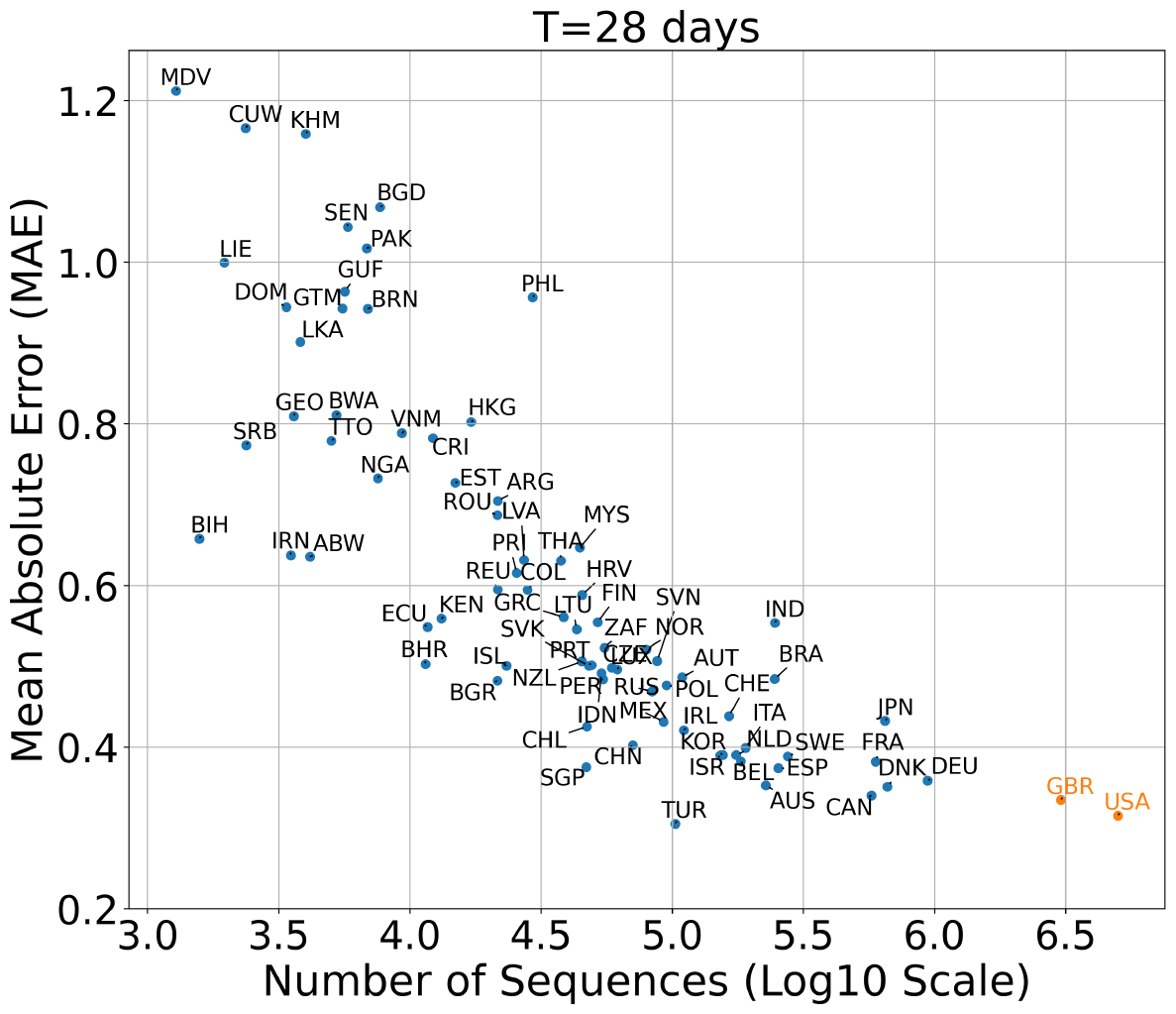
The transformer model trained on data from the US and the UK exhibits remarkable generalization ability when tested on data from many other countries. The figure shows the MAE for the 28-day prediction against the total number of sequences collected before Feb. 26, 2024 in each country. Country codes were indicated around the data points. There is a significant linear correlation between the total number of sequences (on the log10 scale) and the MAE of our model predictions. The Pearson correlation coefficient is *−*0.82, and the p-value is <10^*−*10^.

During model development, we tailored our model specifically to characteristics of the input data and the need to identify lineages that may rise to a high frequency. This includes estimating the target frequency by smoothing the time series to remove noise, using a single-layer transformer to reduce model complexity, and using an exponential weighting function to focus on learning high-frequency lineages (see Methods). To test the effectiveness of these model settings, we performed ablation tests using alternative models where either raw frequencies were used as target frequencies (called ‘Trained with Raw Data’), a two-layer transformer was implemented (called ‘2 Layer Transformer’), or no weighting function was applied (called ‘No Weighted Loss’). We found that none of the alternative models performed better than our original model (Fig. S3), emphasizing our choice of model setting indeed improves overall performance.

### Transformer models outperform Nextstrain predictions

Currently, a widely accepted tool for lineage frequency forecasting is Nextstrain’s multinomial logistic regression (MLR) model (Hadfield et al. 2018; Abousamra et al. 2023; Nextstrain 2023).

Note that the NextStrain MLR model has stringent data criteria, requiring for example that a lineage have at least 5,000 sequences in the past 150 days for a given day of access. This makes it impossible to make early predictions on a newly emerged lineage. In contrast, our model makes predictions on all lineages that have at least 14 days of records in the past, and has no formal requirement for the minimum number of sequences. Therefore, our model is able to make predictions for many more lineages than the NextStrain model and for those lineages that were included in NextStrain model prediction, our model makes predictions much earlier than NextStrain. Nonetheless, we compared our model predictions on the lineages and days where predictions from the Nextstrain MLR model were available.

In general, our model had much better performance across categories (Fig. 5). For example, after we normalized the frequencies of all existing lineages predicted from our models, the MAEs of our model are between 0.28 and 0.36 for 14, 21, and 28-day predictions, only slightly above the noise level. In contrast, the Nextstrain MLR model had MAE greater than 0.5 across all predictions (Fig. 5a). Our transformer model performed particularly well when the predicted frequency was high. That is, it can identify lineages that reached high frequencies, whereas the Nextstrain MLR model performed poorly (Fig. 5b). We also provide visualization examples of 14-day predictions, including 12 lineages from four countries (GBR, USA, CHN, and AUS), in Fig. S4. Overall, these results highlight the superiority of our approach in terms of overall accuracy and stability.

**Figure 5.**
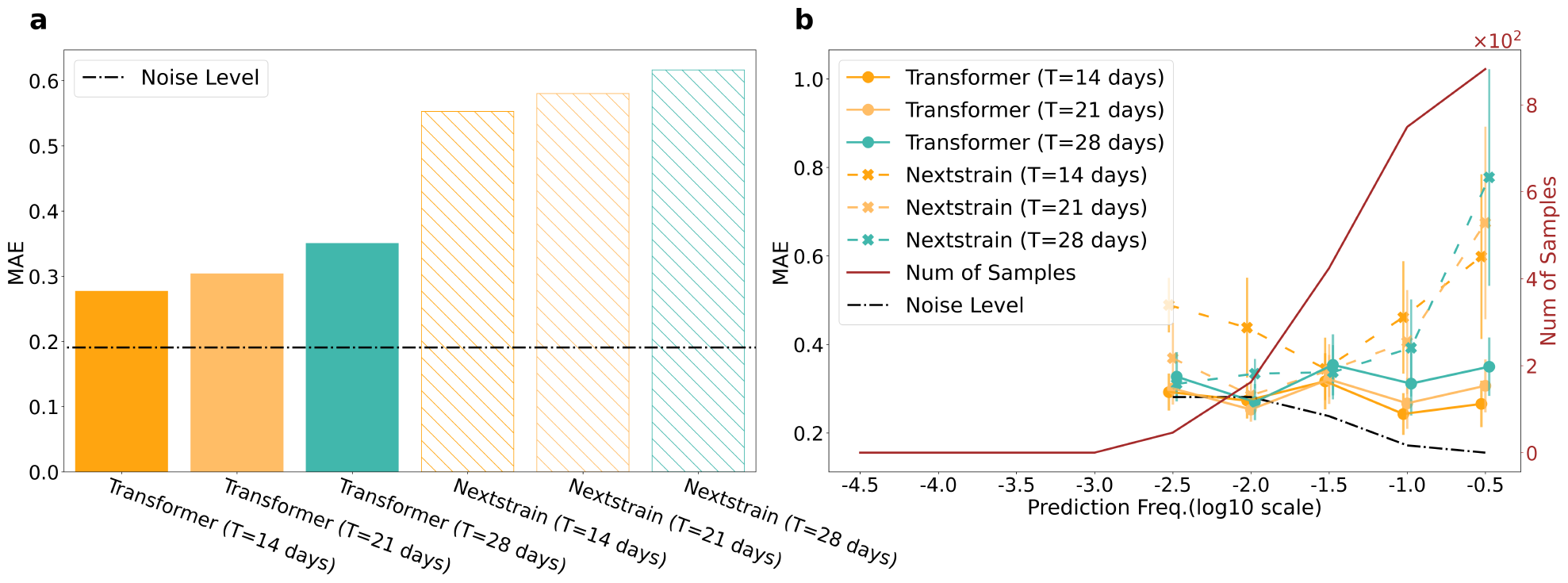
The transformer model outperforms Nextstrain’s Multinomial Logistic Regression (MLR) model. To ensure fair comparison, the MAEs for the transformer model were calculated for countries and dates where the Nextstrain MLR model predictions exist. Only results for 14, 21, and 28 days’ forecasting are shown, because the Nextstrain MLR model forecast a maximum of 30 days into the future. Our models outperform Nextstrain’s MLR model both in terms of overall results **(a)** and stability across different prediction frequencies **(b)**.

## Discussion

Here, we developed a transformer-based model to make accurate forecasts on SARS-CoV-2 lineage frequencies 14 to 60 days into the future. The model excels in robustness, accuracy, and generalization ability across datasets. Its ability to outperform existing models, including the popular Nextstrain multinomial regression model, and adapt to diverse global data sets makes it as a powerful tool for SAR-CoV-2 lineage frequency forecasting and pandemic monitoring.

Although trained only on partial data from the US and the UK, our model demonstrated remarkable generalization ability across datasets from 31 countries in distinct geographic regions across the globe. This is especially important for pandemic monitoring purpose, because it enables early identification of a highly transmissible variant that can appear in any geographic region. In addition, when our model predicts that a lineage may become dominant in multiple countries, it serves as a strong indication that the lineage may become a global variant of concern.

Existing tools for lineage frequency forecasting, such as the multinomial regression approach implemented in the Nextstrain model (Nextstrain 2023; Abousamra et al. 2023), mostly adopt a regression-based approach. This type of approach implements regression on past lineage time series and projects future trajectories based on estimated growth rates. As we demonstrated in this work, these approaches suffer from large noise and biases in the recently collected data points due to reporting delay. In addition, in our previous work (van Dorp et al. 2021), we found that long-time series are needed for regression-based approaches to make reliable predictions due to the uncertainties in the prediction interval. Indeed, the multinomial regression model implemented in Nextstrain has stringent criteria for prediction (e.g., a lineage must have at least 5000 sequences in the past 150 days) to ensure prediction accuracy. In contrast, the transformer-based models we developed here overcome these challenges and make reliable predictions from a minimum of only 14 days of non-zero data in the past 42 days.

Our model’s exceptional performance and ability to generalize, even with a limited training dataset, stem from various strategies specifically designed according to the unique features of the dataset. First, early identification of emerging lineages that eventually become dominant in the population is critical for any lineage forecasting tool. One inherent issue with the data is that lineages that eventually dominated the population are few compared to those that did not, leading to a data imbalance for machine learning model training. By introducing an adaptive loss weighting mechanism to address the data imbalance, we ensured our model prioritizes accurate predictions for rapidly growing lineages that will rise to a high frequency. We demonstrated our model is much better at this task compared to regression-based approaches. Second, to increase the accuracy of long-term prediction, we employed an incremental learning strategy that uses the trained short-term predictions to facilitate long-term predictions. Third, we implemented cross-validation and noise injection techniques (Orvieto et al. 2022) to increase the stability of the model, and this works well for countries with a relatively high sequencing effort. However, the model performance becomes poorer for countries with less sequencing data available (for example, countries with less than 30,000 sequences collected before 2024), suggesting the noises and biases in the datasets from these countries are too large for our current model.

Despite the good performance, there are limitations to our model. First, our model only makes predictions on lineages already existing in the database. As with other forecasting models, it does not predict the origination of new lineages. The origin of a new lineage that transmits efficiently in the population may completely change the dynamics of existing lineages. For example, for the Omicron BA.1 outbreak in late 2021, our model does not make predictions about it before it appeared in the database; however, the model makes accurate predictions of Omicron lineage frequencies once there is enough time series data available. Second, our transformer model was trained on a time series of lineages individually. Although we normalized the predicted frequencies of all existing lineages to 1 for real-time predictions, our model fundamentally only focuses on the dynamics of individual lineage without considering the interactions among co-existing ones. One potential future direction is to develop models to make predictions based on all existing lineages. However, data collected on a single day only lead to one training data point for the model, in this case. Currently, the amount of available data (on the order of a thousand data points) is not sufficient to train such a transformer-based lineage interaction model. Third, our model was trained on data collected from the UK and the US. One potential improvement to the model is to add a country token in the input data stream, train the model using data from more countries, and make country-specific predictions. However, currently, the amount of data from most countries is insufficient to train a model with a country token to make substantial improvements compared to our existing model. As more and more data becomes available, our model can be improved by implementing this strategy. This also points towards the importance of continuing and broadening the genomic surveillance of SARS-CoV-2 lineage evolution.

Overall, our transformer-based model is adept at navigating the challenges posed by substantial noises, biases and missing data inherent in lineage frequency datasets. It not only demonstrates substantially improved forecasting accuracy but also exhibits remarkable generalization ability across numerous countries worldwide. It thus demonstrates that modern machine learning-based approaches represent a promising framework going forward to advance the field of outbreak analysis and epidemiological forecasting.

## Methods

### SARS-CoV-2 data

All available SARS-CoV-2 sequence metadata was downloaded from GISAID (Elbe & Buckland-Merrett 2017) on 2024-02-26. We used the Pango lineage designations provided by GISAID, discarding sequences that lacked Pango or country information. We then summarized the data as the number of records for each combination of collection date, submission date, and country. These counts were used to compute the variant frequencies used in our analysis. In particular, our model was trained and evaluated using datasets from the UK and the US, regions with the highest collection of SARS-CoV-2 genomic sequences. We divided the dataset of all lineages into training and validation sets comprising data up to 2022/12/31, and a testing set with data following this date. This partition resulted in 107,712 entries for training and validation, and 37,707 entries for testing.

### Regression model for lineage frequency forecasting for individual lineages

We assumed that the frequency of a variant lineage would change exponentially, and thus transformed the frequency of each lineage to a log10 scale. We then performed a linear regression on the time series of the log10 lineage frequencies for the past 42 days. To forecast the lineage frequency into the future, we extrapolated lineage frequency based on the parameters from the linear regression. When the extrapolated frequencies are greater than 100%, we assume the predicted frequency is 100%.

### Data processing for the transformer models

To improve the data quality, we first removed isolated data points during data pre-processing. Specifically, a data point is identified as potentially anomalous and removed if there are five or more days without any records within a seven-day period centered on that point. The input of the machine learning model is constructed as a 5*n* dimensional vector, where *n* is the number of input days. For each day, we incorporate five features: the frequency of the specific lineage, the number of sequences of the lineage, the total number of sequences on that day, the number of days elapsed since 2020-01-01, and the elapsed days since the lineage’s first recorded collection day. For the missing records, we use a fixed negative token as a placeholder. By using special tokens, transformers can effectively manage missing or masked data, making them robust tools for a wide range of sequence-related tasks. In particular, we treat each one-day feature as a discrete element (referred to as a token or patch) for input processing. Thus, the patch size is 1 *×* 5. The model’s output is the predicted frequency of the lineage for a future time point, specified as *T* days ahead.

Given the presence of noise and missing data in our dataset, we employ a 1-D smoothing spline following interpolation to smooth the ground truth, which we then use as our labels.

### Machine learning framework

We employed a single-layer transformer with a linear input layer preceding it and a linear output layer following it. As shown in Fig. 1b, the transformer contains a self-attention layer, followed by an MLP. The overall model architecture is illustrated in Fig. 1b. In particular, the model uses embedding dimensions of 8 and 2 attention heads without dropouts. There is a layer normalization between the transformer and the input/output linear layer, separately. For the position embedding, we employed the fixed sin-cos embedding described in the original paper (Vaswani et al. 2017). Here, the primary challenge in our problem lies in the presence of significant data noise. Our primary objective is not to extract deeply hierarchical or intricate semantic information, but rather to uncover valuable signals within this noisy data. Given the relatively low amount of data for model training, our foremost considerations are robustness, simplicity, and resistance to overfitting. Consequently, we have deliberately opted for a shallower network architecture.

While the model is easier to learn the short-term prediction, we used an incremental method to enhance longterm predictions by including short-term predictions. As shown in the dotted line part of Fig. 1b, in particular, we trained a model that can predict 14 days’ future first. Then, the 14-day predictions (ensemble of five models’ results from 5-fold cross-validation) are concatenated with the input feature. Additionally, we introduce a shortcut connection that adds the short-term prediction to the model’s output to calculate the final long-term predictions. This approach combines short-term insights with long-term forecasting, resulting in improved accuracy.

For the purpose of training, validating, and testing our model, we partition the data as of 2023-01-01. Data recorded prior to this date are allocated for model training and validation with 5-fold cross-validation. Then, the best-performed models will be ensembled and tested on data subsequent to this date as the testing data set. Our model is trained on combined datasets from the United States (USA) and Great Britain (GBR). Moreover, we utilize noise injection (Orvieto et al. 2022) technique to improve the model generalization ability, which perturbs the model parameters by random Gaussian noise (zero means and 1*e−* 4 std) in each training iteration. This prevents the model from converging to sharp local minima.

To address the challenge of data imbalance inherent in our dataset, we have employed an approach wherein we apply an exponential function to weight the loss function during model training. This weighting strategy takes into account the varying frequencies of the target day, assigning higher weights to datapoints corresponding to high-frequency labels. By doing so, the model places greater emphasis on accurately predicting the high-frequency variants, which are often of paramount importance in epidemiological and public health contexts. This adaptive weighting mechanism ensures that the model focuses on effectively capturing the dynamics of the most prevalent variants, ultimately leading to improved overall forecasting performance and a more balanced predictive outcome. The overall loss function combines *l*_1_ and *l*_2_ loss, with an exponential weighting scheme. This loss function is designed to give more emphasis to lineages with higher frequency. The loss function can be mathematically represented as:

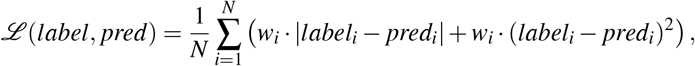

where *N* represents the number of samples, *label*_*i*_ is the true label of the *i*-th sample, *pred*_*i*_ is the model’s prediction for the *i*-th sample, and *w*_*i*_ is the weight associated with the *i*-th sample, which is computed as 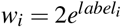.

To train the model, we employed AdamW (Loshchilov & Hutter 2018) optimizer with momentum parameters *β*_1_ = 0.9, *β*_2_ = 0.999 and a weight decay of 0.05. The initial learning rate is set to be 1 *×* 10^*−*3^, and we modify the learning rate with a cosine annealing (Loshchilov & Hutter 2016). We set the batch size to 256 and train the model for 1000 epochs. We implemented our models in Pytorch and trained them on 1 NVIDIA Tesla V100 GPU.

## Data Availability

All data produced in the present study are available upon reasonable request to the authors.

## Acknowledgements

Research presented in this article was supported by the Laboratory Directed Research and Development program of Los Alamos National Laboratory under project number 20230830ER. This research used resources provided by the Darwin testbed at Los Alamos National Laboratory (LANL) which is funded by the Computational Systems and Software Environments subprogram of LANL’s Advanced Simulation and Computing program (NNSA/DOE).

---

**Figure S1:**
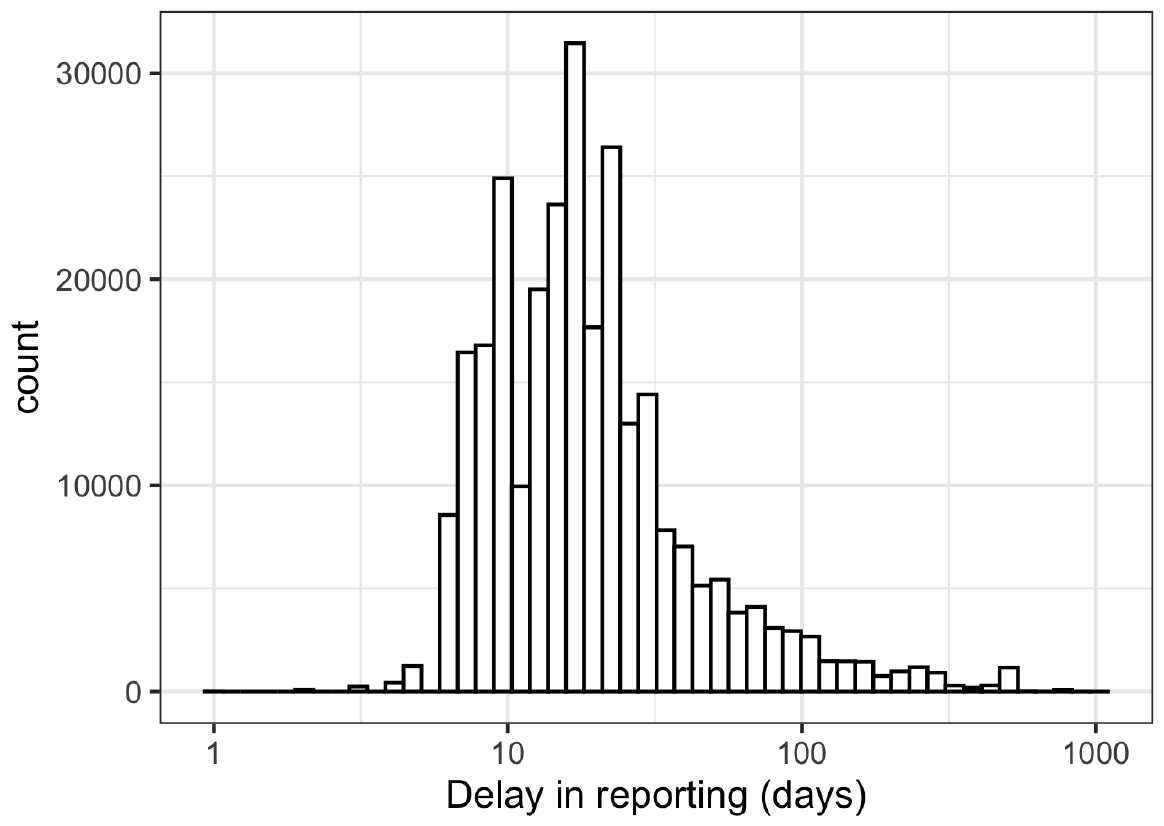
Distribution of the reporting delays. The difference between the date of sample collection and the date of reporting, calculated for data from the UK and the US.

**Figure S2:**
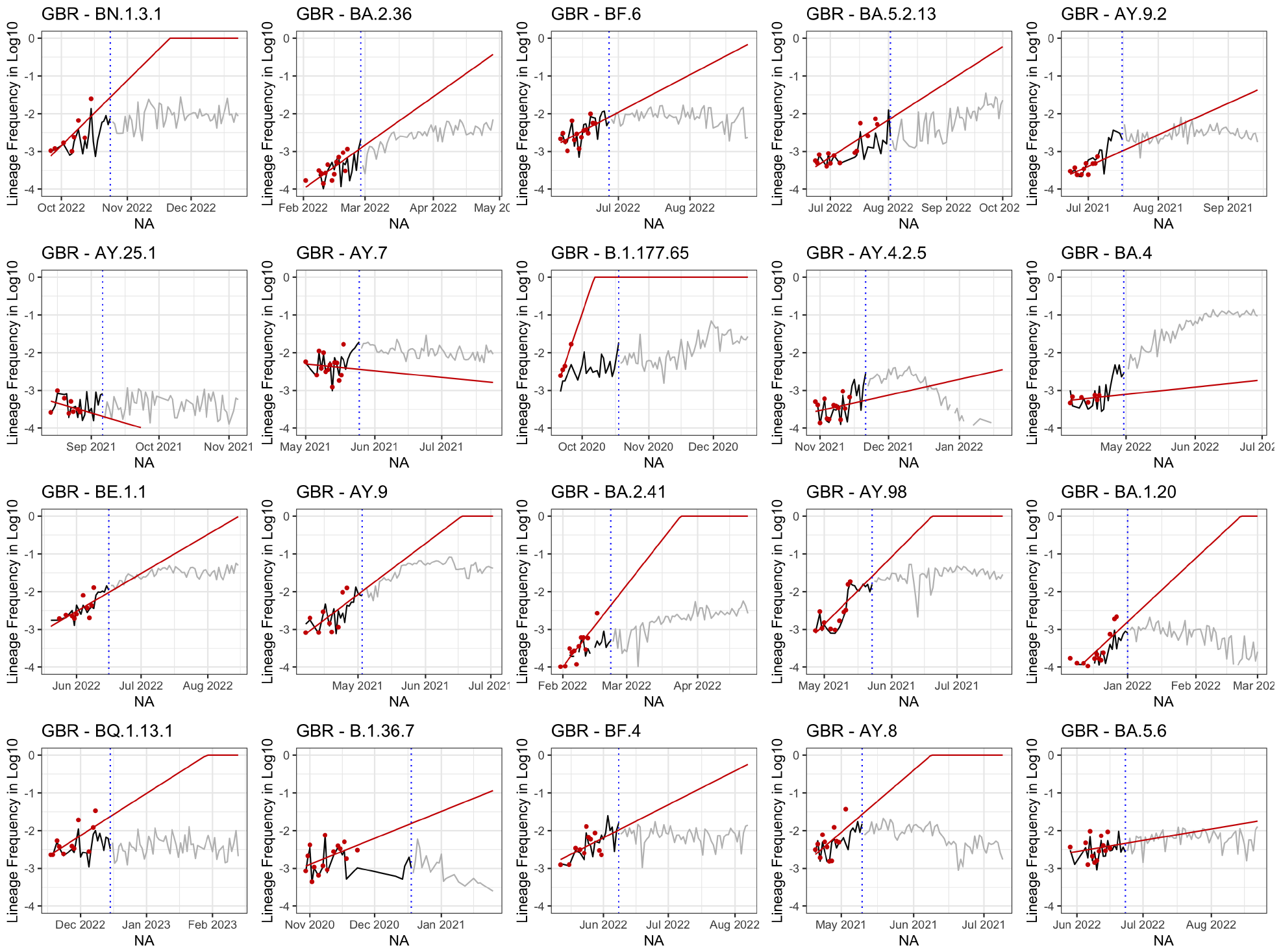
A linear-regression-based forecasting model does not predict future trajectories accurately. Poor predictions are due to noise and biases from the intrinsic process and reporting delay. Shown are 20 randomly selected lineages in the UK. A linear regression (red lines) is performed on the lineage frequency time series (in log10; red dots) that were calculated based on the access date marked by the blue dotted lines. The future projections are based on the regression line. If the projected frequencies are greater than 100%, they are assumed to be 100%. Black and grey lines represent the final lineage frequencies before and after the assumed access date, respectively.

**Figure S3:**
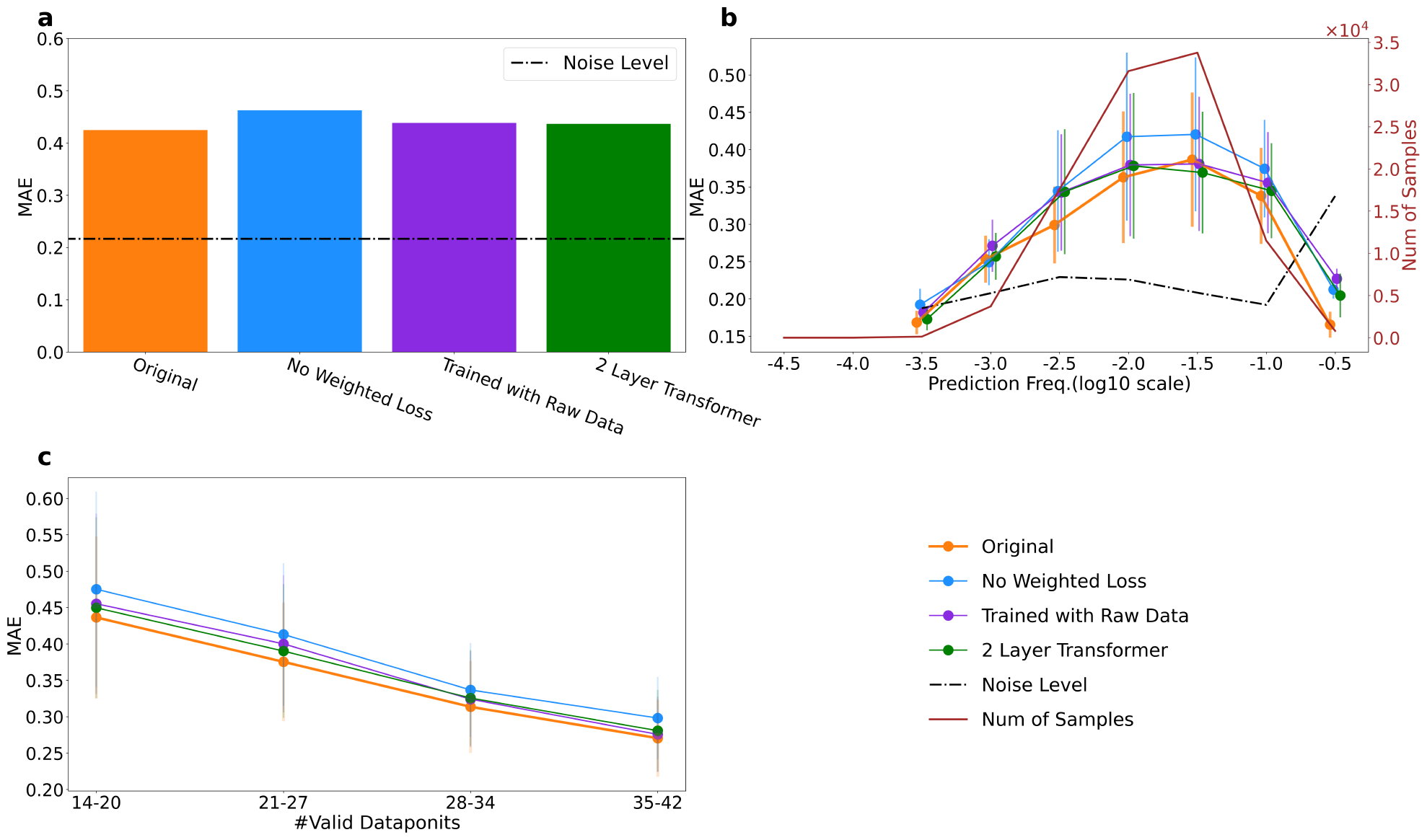
Ablation test results of our model (‘Original’) compared with three other alternative model variants on all countries for 14-day prediction. For the ‘No Weighted Loss’ model, the loss function is calculated as the MAE without the exponential weighting function applied. In the ‘Trained with Raw Data’ model, raw frequencies were used as target frequencies (instead of using a smoothing spline). In the ‘2 Layer Transformer’ model, a two-layer transformer was implemented. All other settings of each model variant are the same as the original model. **(a)** Overall performance of the four models. **(b)** MAEs categorized by the predicted frequency from each model. Our model demonstrates better overall performance across various outputs. Especially for high-frequency results that are more worthy of concern, our models are much more effective. **(c)** MAEs categorized by the number of non-zero data points in the input time series.

**Figure S4:**
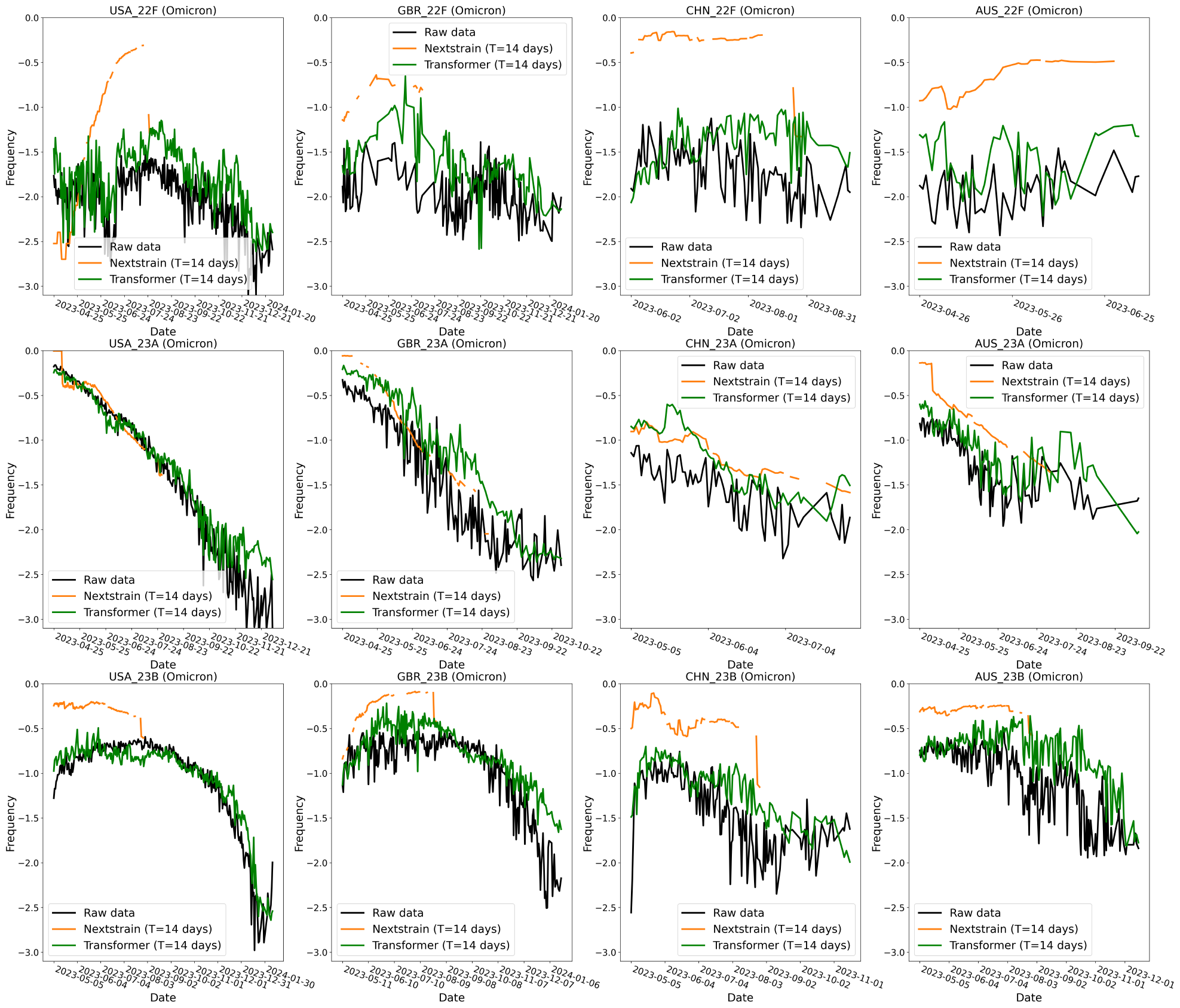
Examples of predictions for 12 clades from our model and the Nextstrain MLR model. The transformer model clearly outperforms the Nextstrain MLR model. Comparisons show raw frequency data (black) and predictions from the transformer model (green) and the Nextstrain MLR model (orange). Lineages were clustered into Nextstrain clades based on CoVariants (2022). Visualizations start on the day that Nextstrain has available predictions.

